# Cytosolic PCNA interacts with S100A8 and controls an inflammatory subset of neutrophils in COVID-19

**DOI:** 10.1101/2022.10.12.22280984

**Authors:** Rodrigo de Oliveira Formiga, Lucie Pesenti, Maha Zohra Ladjemi, Philippe Frachet, Muriel Andrieu, Souganya Many, Vaarany Karunanithy, Karine Bailly, Théo Dhôte, Manon Castel, Christophe Rousseau, Marick Starick, Edroaldo Lummertz da Rocha, Emilia Puig Lombardi, Vanessa Granger, Sylvie Chollet-Martin, Luc De Chaisemartin, Luc Mouthon, Fernando Spiller, Anne Hosmalin, Margarita Hurtado-Nedelec, Clémence Martin, Frédéric Pène, Pierre-Regis Burgel, Léa Tourneur, Véronique Witko-Sarsat

**Affiliations:** Université Paris Cité, INSERM U1016, CNRS 8104, Institut Cochin, Paris, France; Laboratory of Immunobiology, Department of Pharmacology, Federal University of Santa Catarina (UFSC), Florianopolis, Santa Catarina, Brazil; Université Grenoble Alpes, CEA, CNRS, IBS, Grenoble, France; Bioinformatics Core Platform, INSERM UMR1163, Imagine Institute, Université Paris Cité, Paris, France; APHP, Immunology department, Auto-immunity, hypersensitivities and biologicals, Bichat Hospital, Paris, France; Paris-Saclay University, INSERM, Inflammation, microbiome and immunosurveillance, Châtenay-Malabry, France; Department of Internal Medicine, AP-HP, Cochin Hospital, Paris, France; Department of Immunology and Hematology, UF Immune Dysfunctions, HUPNVS, Bichat Hospital, Paris, France; Department of Pneumology, AP-HP, Cochin Hospital, Paris, France; Department of Intensive Medicine and Reanimation, AP-HP, Cochin Hospital, Paris, France

**Keywords:** Neutrophils, PCNA, S100A8, COVID-19

## Abstract

Neutrophils are key players in the hyperinflammatory response upon SARS-CoV-2 infection. We have previously described that cytosolic proliferating cell nuclear antigen (PCNA) controls neutrophil survival and NADPH oxidase-dependent ROS production. We here show that both PCNA and S100A8 expression and interaction were elevated in neutrophils from patients with COVID-19 compared to healthy donors and this was correlated with disease severity. Increased PCNA expression was accompanied by a decreased apoptosis and increased NADPH-oxidase activity in neutrophils from COVID-19 patients compared to healthy donors. These effects, as well as the interaction between PCNA and S100A8, were potently counteracted by T2 amino alcohol (T2AA), a PCNA inhibitor, demonstrating that the PCNA scaffold orchestrated neutrophil activation. Notably, the interaction between PCNA-S100A8 was more intense in the CD16^high^-CD62L^low^ activated neutrophil subset. We propose that PCNA-S100A8 complex acts as potential driver for neutrophil dysregulation in COVID-19 and show for the first time that the PCNA scaffold is a decisive component of both neutrophil activation and heterogeneity.

## Introduction

Neutrophils are key players in the innate immune response and form the first line of defense against infectious agents, as they employ well-characterized anti-microbial strategies, such as phagocytosis or release of reactive oxygen species (ROS)^1^, neutrophil extracellular traps (NETs)^2^ and other pro-inflammatory mediators.^3,4^ Recent unexpected biological features, such as their phenotypic and functional heterogeneity,^5^ their extended lifespan under certain pathological conditions have highlighted their crucial role in controlling adaptive immune responses.^6–8^ Increased neutrophil counts and newly identified subsets with characteristics of immaturity, immunosuppression, activation, and metabolic rewiring in both circulation and infiltrated into the respiratory tract may be pivotal clinical aspects of severe Coronavirus Disease (COVID)-19 caused by severe acute respiratory syndrome coronavirus 2 (SARS-CoV-2).^8^ Upon SARS-CoV-2 infection, a wide spectrum of clinical outcomes may appear, ranging from self-limited asymptomatic^9^ or mild disease to severe cases, the last marked by thromboembolic events,^10^ dysregulated immune response,^11^ and respiratory distress and multiorgan failure.^8,12–15^

In COVID-19 the circulating levels of calprotectin, also known as S100A8/A9, are highly abundant and are associated with immune imbalance and expansion of aberrant neutrophil subpopulations that correlated with disease severity.^16–17^ Indeed, calprotectin is a potential biomarker in a wide number of inflammatory conditions in which neutrophils are involved, such as autoimmune vasculitis,^18^ arthritis,^19^ cardiovascular, and metabolic disorders. The S100A8/A9 is constitutively expressed in neutrophils and monocytes, a Ca2+ sensor, participates in cytoskeleton rearrangement and arachidonic acid metabolism.^20^ Neutrophil secretion of S100A8/A9 might act as a extracellular signaling mediator.^21^

To gain insights into the molecular mechanisms involved in neutrophil dysregulation and reprogramming observed in COVID-19 patients, we examined the expression and function of the proliferating cell nuclear antigen (PCNA), which we previously described as a key regulator of neutrophil fate.^22,23^ PCNA is a scaffolding protein expressed in the nucleus of proliferating cells where it controls DNA replication and repair.^24,25^ By contrast, in neutrophils, which are non-proliferative cells, PCNA is exclusively cytoplasmic^26^ and interacts with procaspases to promote neutrophil survival.^22,27^ Notably, T2 amino alcohol (T2AA)^28,29^, a small molecule inhibitor that specifically binds to PCNA promotes the resolution of inflammation in a model of murine colitis. Proteomic analysis of neutrophil PCNA interactome identified a diversity of proteins involved in different functions, including cytoskeletal regulation, metabolism,^30^ and NADPH oxidase. Indeed, PCNA interacts directly with p47^phox^, a NADPH oxidase subunit, to control the oxidative burst.^31^ Likewise, calprotectin promotes NADPH oxidase complex assembly and ROS generation through direct interactions between S100A8 and the cytosolic subunit p67^phox32^ and between S100A9 and the cytochrome b558 protein, gp91^phox^.^33^

The aim of this study is to examine whether and how the cytosolic proteins PCNA and S100A8/A9 may be involved in the dysregulated function of neutrophils in COVID-19 patients of various severity. We herein identify a pivotal role of cytosolic PCNA in promoting survival and NADPH oxidase-dependent ROS production in neutrophils, notably through its enhanced interaction with the S100A8 protein. Most strikingly, this PCNA-S100A8 interaction in neutrophil cytosol was found in a particular neutrophil subset and was related to COVID-19 severity.

## Results

### Increased PCNA expression is associated with increased survival in neutrophils from COVID-19 patients

Western blot analysis of PCNA in neutrophils from HD and COVID-19 patients showed an increase in PCNA cytosolic expression correlating with disease severity with significant differences between critical patients and HD, as well as severe patients and HD. However, no significant changes were found between HD/moderate or severe/critical patients (Figures 1A-B). Increased PCNA expression in neutrophils from both severe and critical COVID-19 patients was confirmed by immunofluorescence labelling of PCNA that displayed a cytoplasmic pattern (Figures 1C-D). Re-analysis of publicly available scRNA-seq data^16,41^ pointed to no changes in PCNA mRNA in circulating or respiratory tract infiltrating neutrophils in severe COVID-19 patients (Supplemental Figure 2). Notably, the PCNA cytosolic expression was correlated with circulating NETs (Figures 1E-F), neutrophil elastase (Figures 1G-H) and calprotectin levels (Figures 1I-J) strongly suggesting that cytosolic PCNA was pivotal in neutrophil activation state and was reminiscent of systemic inflammation.

**Figure 1.**
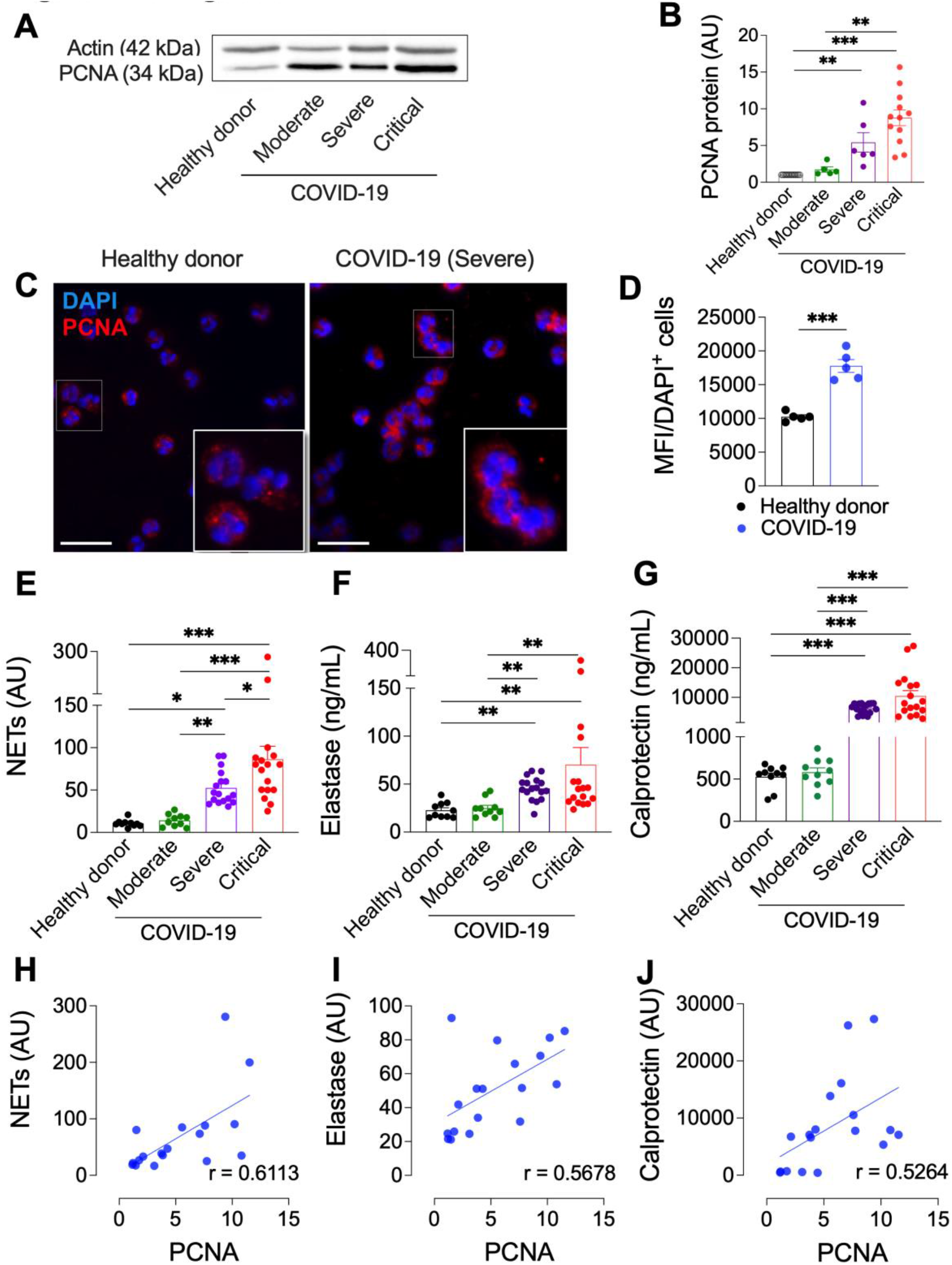
PCNA is overexpressed in the cytosol of neutrophils and is associated with COVID-19 severity. **(A-B)** Western blot analysis of PCNA expression in neutrophils cytosols from HD (n = 17), moderate (n = 5), severe (n = 6), and critical (n = 12) COVID-19 patients. **(A)** Representative experiment. **(B)** Quantification of Western blots. Data are expressed as fold change in reference to PCNA expression in HD. **(C-D)** Neutrophils from HD (n = 5), severe or critical COVID-19 patients (n = 5) were labeled with a rabbit polyclonal anti-PCNA antibody and with DAPI to visualize nuclei using the Widefield Zeiss Observer Z1 microscope (magnification x 100). **(C)** Representative experiment of indirect immunofluorescent staining of PCNA showing its cytosolic localization (red) and the nuclei (blue). **(D)** Quantification of the PCNA immunolabelling fluorescence. Photos were captured from at least 3 different fields for a given donor and each point corresponds to the mean of a different donor. **(E-G)** Soluble inflammation markers were measured: **(E)** NETs, as MPO-DNA complexes; **(F)** Elastase and **(G)** S100A8/A9 (calprotectin), by ELISA in serum from HD (n = 12) moderate (n = 7) severe (n = 18) and critical (n = 17) COVID-19 patients. **(H-J)** Correlation between normalized cytosolic PCNA expression measured by Western blot in neutrophils from patients with all levels of COVID-19 severity (as in B) and the circulating concentration of **(H)** NETs **(I)** Elastase and **(J)** Calprotectin. Pearson correlation coefficient are indicated and the associated probability is: **p = 0.007, *p = 0.014 and *p = 0.024, respectively. Graph symbols represent individual donors. In B, D, E, F, and G data are presented as mean ± SEM. Data was analyzed by one way-ANOVA followed by Tukey or Dunnett post-test or by Student’s t-test when two individual groups were compared (*p<0.05; **p<0.01; ***p<0.001).

We next evaluated the ability of neutrophils from COVID-19 patients to undergo spontaneous apoptosis after a 16h *in vitro* incubation by measuring annexin-V binding on externalized phosphatidylserine. As expected, neutrophils from severe and critical COVID-19 patients were significantly more viable, less apoptotic, and less necrotic (evaluated by 7AAD labelling) than HD neutrophils (Figure 2A-C). Notably, T2AA, a PCNA inhibitor,^28,29^ significantly decreased the survival of COVID-19 neutrophils by increasing apoptosis (Figure 2A-C). This increased survival was confirmed by a significantly stronger DiOC6 labelling thereby indicating a preserved mitochondrial membrane potential (Δψm) during overnight incubation as compared to HD. This sustained Δψm was reversed by disrupting the PCNA scaffold by T2AA (Figure 2D), again highlighting the key role of PCNA in survival of COVID-19 neutrophils.

**Figure 2.**
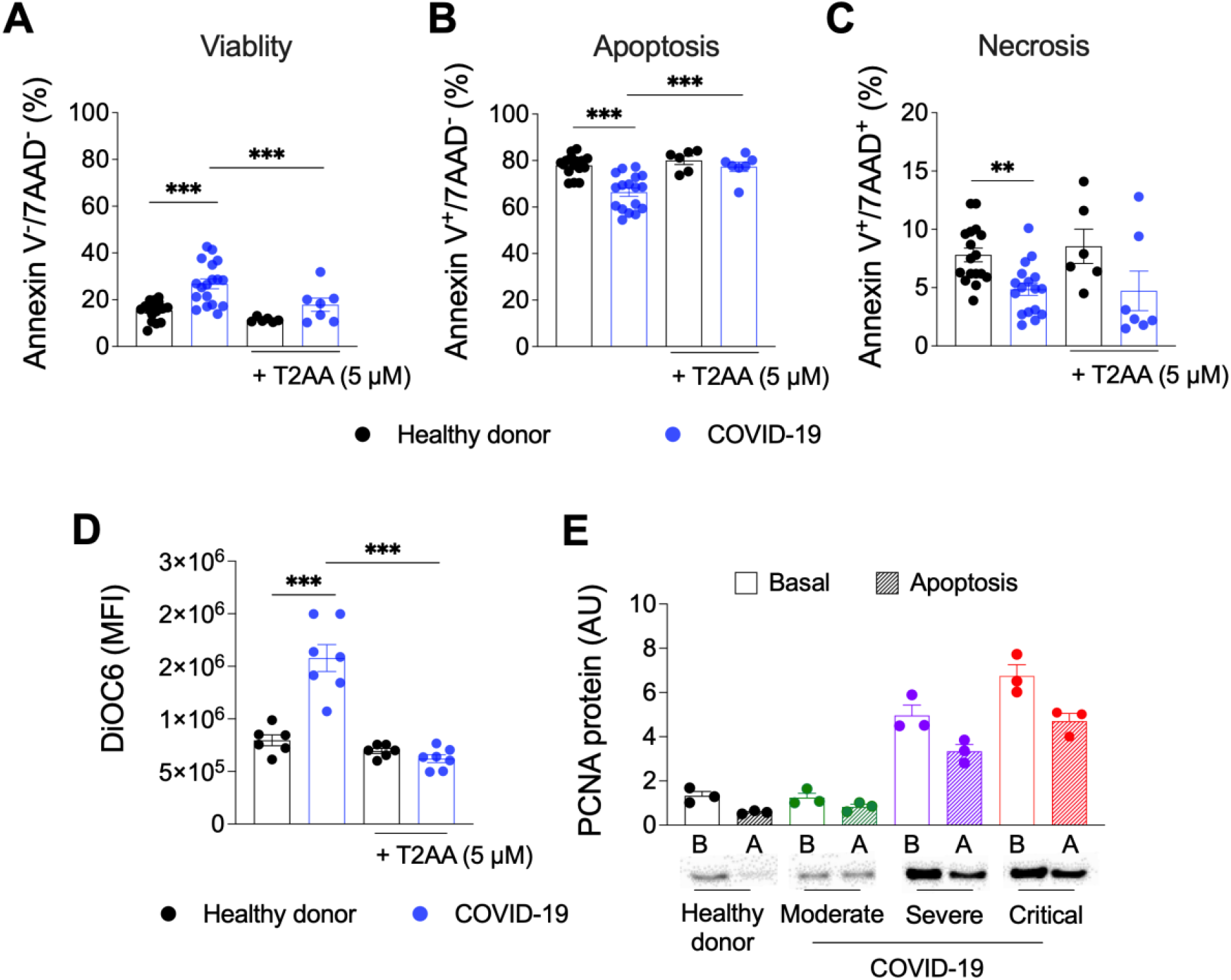
Increased neutrophils survival in COVID-19 is reversed by the PCNA inhibitor T2AA. **(A-D)** Spontaneous apoptosis was induced by overnight incubation of neutrophils from HD (n = 10) severe or critical COVID-19 patients (n = 10) at 37°C, 5% CO_2_ for 16 h in the presence or absence of the PCNA inhibitor T2AA (5 μM) **(A)** Percentage of viable neutrophils with annexin-V^+^/7-AAD^+^ labelling. **(B)** Percentage of apoptotic neutrophils with annexin-V^+^/7-AAD^−^ labelling. **(C)** Percentage of necrotic neutrophils with annexin-V^+^/7-AAD^+^ labelling. **(D)** Flow cytometry measurement of DIOC6 labelling to assess mitochondria integrity. MFI of DiOC6 staining is indicative of intact mitochondrial membrane potential and is decreased upon apoptosis. **(E)** PCNA expression measured by Western blot in cytosols from freshly isolated neutrophils at basal state [B] or following a 16h-incubation triggering spontaneous apoptosis [A]. PCNA abundance was quantified by densitometry and normalized using Ponceau red staining. Data are the mean of three independent experiments carried out with neutrophils from HD and COVID-19 patients with different disease severity (moderate, severe, or critical). In A, B, C and D, graph symbols represent individual donors. Data are presented as mean ± SEM and were analyzed by one way-ANOVA followed by Tukey or Dunnett post-test or by Student’s t-test when two individual groups were compared (***p<0.001).

In accordance with the notion that cytosolic PCNA inhibits apoptosis,^22^ Western blot analysis confirmed that PCNA expression was significantly decreased after apoptosis in HD neutrophils. By contrast, COVID-19 neutrophils maintained high amounts of PCNA even after spontaneous apoptosis and this was particularly pronounced in critically ill patients who display the highest level of PCNA (Figure 2E).

### Cytosolic PCNA controls sustained NADPH oxidase-dependent ROS production in neutrophils from COVID-19 patients

We next examined NADPH oxidase-dependent ROS production to evaluate the activation state of COVID-19 neutrophils. No significant difference in ROS production was found between neutrophils from COVID-19 patients with a moderate severity compared to HD at basal state or after *in vitro* activation by PMA, opsonized zymosan or f-MLF. By contrast, neutrophils from severe and critical COVID-19 patients had a significant increase in basal ROS production compared to HD showing that they were activated in the absence of *in vitro* stimulus (Figure 3A). ROS production was significantly increased after PMA, opsonized zymosan, or f-MLF stimulation in severe COVID-19 patients as compared with HD, suggesting that this increased ROS production was associated with the severity of the disease to some extent. Notably, inhibition of the PCNA scaffold by T2AA resulted in a significant decrease in either basal (Figure 3A) or stimulated ROS production (Figure 3B-D) in COVID-19 neutrophils whatever the severity of the disease. Accordingly, T2AA decreased NADPH-oxidase dependent ROS production at higher concentrations in COVID-19 compared to HD neutrophils (Supplemental Figure 3). Taken together, our data suggest that PCNA sustains NADPH oxidase-dependent ROS production in COVID-19 neutrophils.

**Figure 3.**
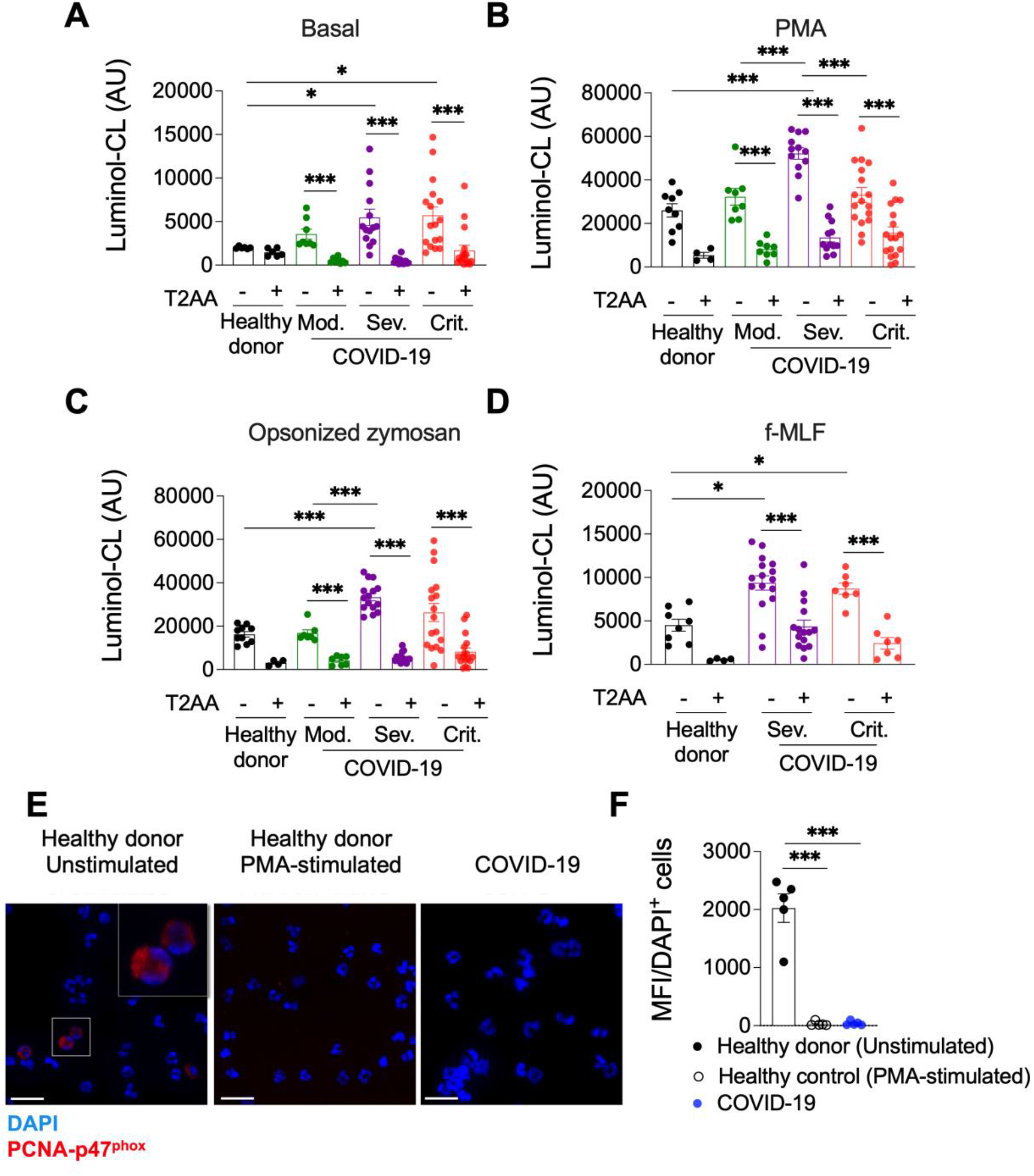
PCNA regulates NADPH oxidase-dependent ROS production in COVID-19. NADPH oxidase activation was assessed by using luminol-enhanced chemiluminescence (CL) that measured ROS production in isolated neutrophils from HD (n = 7), moderate (n = 8), severe (n = 14) and critical (n = 17) COVID-19 patients under **(A)** basal or following stimulation by **(B)** PMA (0.1 μg/mL), **(C)** opsonized zymosan (0.5 mg/mL) or **(D)** f-MLF (1 μM). These measurements were performed in the absence or in the presence of the PCNA inhibitor, T2AA (5 μM). CL was recorded at 37°C in a Tristar™ luminometer over time and data were expressed as arbitrary units (AU) at the peak of ROS production. **(E-F)** Assessment of the PCNA-p47^phox^ interaction in neutrophils. The Duolink^®^ proximity ligation assay (PLA) was performed on isolated neutrophils from HD (n=5) or severe or critical COVID-19 patients (n=5). Neutrophils were incubated in the absence or in the presence of PMA for 10 min to trigger NADPH oxidase activation. **(E)** Representative experiment of PLA staining using rabbit polyclonal anti-PCNA antibody associated with mouse monoclonal anti-p47^phox^ antibody showing the interaction of PCNA and p47^phox^ (red) and the nuclei labelled by DAPI (blue) and visualized using the Widefield Zeiss Observer Z1 microscope (magnification x 100). **(F)** Quantification of the PCNA-p47^phox^ proximity-induced fluorescence. Photos were captured from at least 3 different fields for a given donor and MFI was analyzed, each point corresponds to the mean of a different donors. Graph symbols represent individual donors. Data are presented as mean ± SEM. Data was analyzed by one way-ANOVA followed by Tukey or Dunnett post-test or by Student’s t-test when two individual groups were compared (*p<0.05; ***p<0.001).

We previously showed that PCNA can directly bind to the cytosolic p47^phox^ protein, acting as a chaperone to promote NADPH oxidase activation.^31^ We here confirmed that the PCNA-p47^phox^ association evaluated by a proximity ligation assay was detected in HD neutrophils at basal state but was absent after PMA activation as previously reported.^37^ By contrast, the PCNA-p47^phox^ association was undetectable in neutrophils from severe COVID-19 patients (Figure 2E-F) strongly supporting the notion that they were activated at the basal state.

### S100A8 is a new partner of PCNA which is overexpressed in neutrophils cytosols from COVID-19 patients

S100A8 or S100A9 have been described to promote NADPH oxidase activity through their association with cytosolic NADPH oxidase components.^32,33^ Since calprotectin serum levels are positively correlated with neutrophil activation and COVID-19 severity,^16,42^ we next tested whether S100A8 or S100A9 could be part of the PCNA scaffold thereby supporting neutrophil dysregulation in COVID-19. Surface plasmon resonance (SPR) was carried out to examine whether PCNA could directly interact with either S100A8 or S100A9 or with the dimeric S100A8/A9 calprotectin. Our data unambiguously showed that there was a direct S100A8 association with immobilized PCNA in a concentration-dependent manner (0.1-2μM) with an equilibrium dissociation constant (K_D_) value within the micromolar range (Figure 4A; Table 2). By contrast, S100A9 and the S100A8/A9 heterodimer failed to associate with PCNA when assayed under the same experimental settings (Figure 4B; Table 2). We confirmed, using a proximity ligation assay, that PCNA and S100A8 were associated within the cytoplasm under basal conditions in neutrophils from both HD and COVID-19 patients but the PCNA-S100A8 proximity-induced fluorescence intensity was significantly increased in the latter. Notably, T2AA treatment, by interfering with the PCNA scaffold, significantly blunted the PCNA-S100A8 interaction (Figure 4C-D). Pertinently, using Western blot analysis, we observed an increased S100A8 cytosolic levels in COVID-19 neutrophils according to disease severity (Figure 4 E-F), showing positive correlation with PCNA expression (Figure 4G). S100A8 immunofluorescence in neutrophils from HD and severe COVID patients confirmed its cytoplasmic localization (Figure 4H). Increased fluorescence intensity consistent with increased S100A8 expression was observed in COVID-19 neutrophils compared to those of HD (Figure 4I). Notably, analysis of publicly available scRNA-seq data^16,41^ showed that S100A8 and S100A9 mRNA percentage and average of expression were altered in COVID-19 peripheral neutrophils, and high S100A8 mRNA levels were detected in respiratory tract infiltrating neutrophils, showing higher expression levels in critical over moderate patients (Supplemental Figures 2).

**Table 2.** Kinetics and affinity of PCNA interaction with S100A8

| PCNA/S100A8 |  |  |  |  |  |  |
| --- | --- | --- | --- | --- | --- | --- |
| Immobilized<br>ligand/Soluble<br>analyte | $K_{a1}$<br>($M^{-1} s^{-1}$) | $K_{d1}$<br>($s^{-1}$) | $K_{a2}$<br>($M^{-1} s^{-1}$) | $K_{d2}$ ($s^{-1}$) | $K_D$ (M) | $\chi^2$ |
| Heterogenous<br>ligand model | 2199 | 0.1035 | 138.1 | 0.0034 | $4.7 \times 10^{-5}$ ($K_{D1}$)<br>$2.49 \times 10^{-5}$ ($K_{D2}$) | 7.6 |
| 1:1 model | 158.5 | 0.00568 | NA | NA | $3.58 \times 10^{-5}$ | 16.6 |

Binding of S100A8 (0.1-2 $\mu M$ ) was measured as described in Materials and Methods section. Values were calculated for 1:1 and heterogeneous ligand models. The heterogeneous ligand model accounts for two different binding sites on the immobilized PCNA, and the two corresponding dissociation constants ( $K_{D1}$ and $K_{D2}$ ) were determined. NA = not applicable.

**Figure 4.**
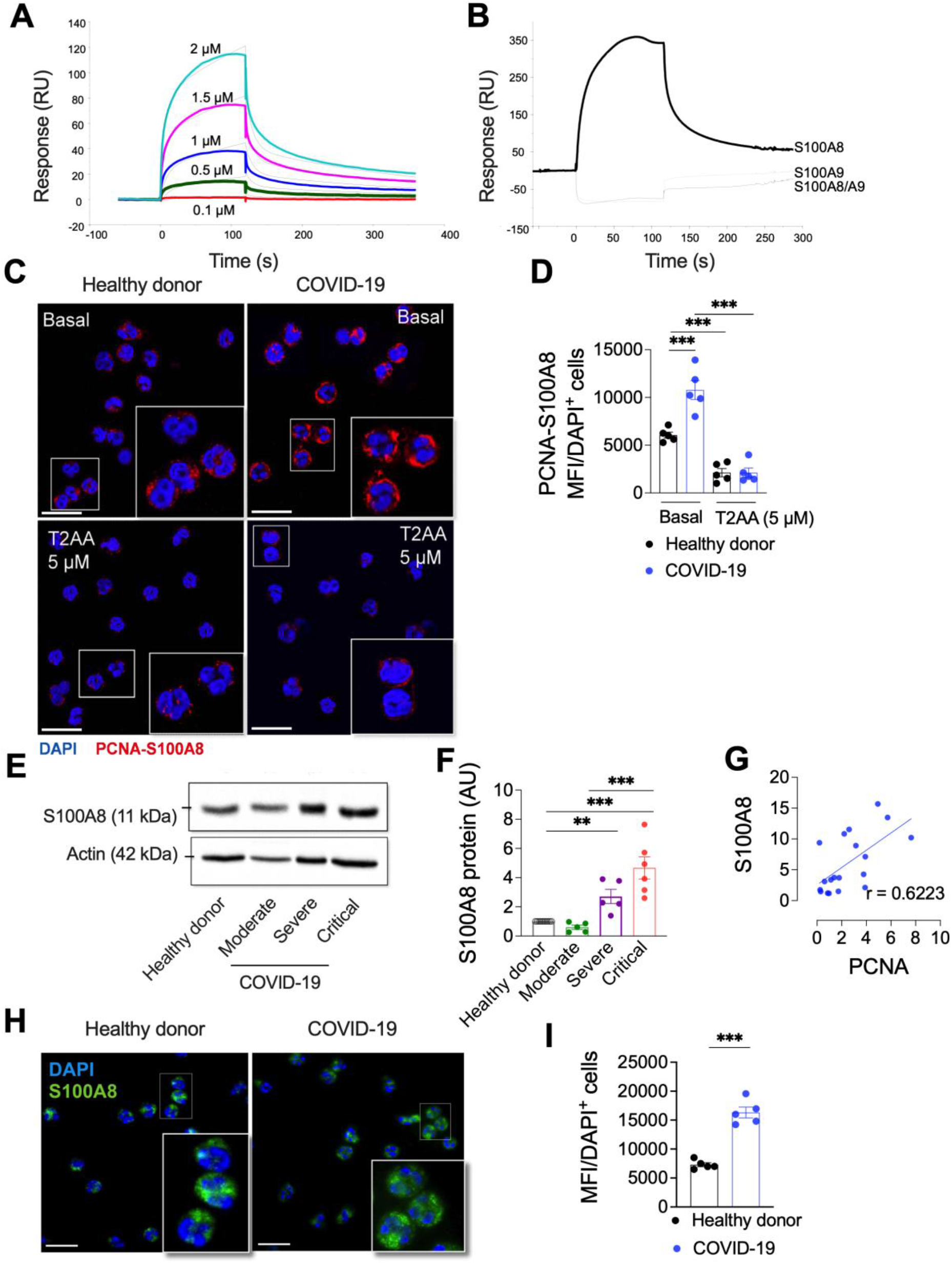
S100A8 is a partner of PCNA overexpressed in neutrophils cytosols from COVID-19 patients. **(A)** Data represent overlays of sensorgrams resulting from injection of human recombinant S100A8 at different concentrations (0.1 – 2 μM). Fits with a statistic χ2 value 7.6 are shown as dotted lines (RU: response units). SPR experiments were performed on a Biacore T200 apparatus. Kinetic values determination is reported in Table 2. Interaction was predicted under a standard curve composed of baseline, association, equilibrium, and dissociation phases. **(B)** SPR measurements of human recombinant S100A8, S100A9 or S100A8/A9 (2 μM) binding on immobilized PCNA. **(C)** Representative experiment of the assessment of the PCNA-S100A8 interaction in isolated neutrophils using the Duolink^®^ proximity ligation assay (PLA). PCNA-S100A8 proximity-induced fluorescence was detected (red) using a rabbit polyclonal anti-PCNA and a mouse monoclonal anti-S100A8. Prior to experiment, neutrophils were incubated in the absence or presence of T2AA (5 μM) for 30 min. DAPI was used for nuclear staining (blue) Cells were visualized using the SR Leica SP8X STED Flim confocal microscope (magnification ×100) **(D)** Quantification of the PCNA-S100A8 proximity-induced fluorescence. Photos were captured from at least 3 different fields for a given donor and MFI was analyzed, each point corresponds to the mean of a different donors (E) Western blot analysis of S100A8 expression in neutrophils cytosols from HD (n = 17), moderate (n = 5), severe (n = 6), and critical (n = 6) COVID-19 patients. **(F)** Quantification of S100A8 abundance by densitometry. Western blot analysis of actin was performed on the same membranes and used as a loading control for the normalization of PCNA expression. **(G)** Correlation between normalized PCNA and S100A8 expressions measured by Western blot in neutrophils from patients with all levels of COVID-19 severity. Pearson correlation coefficient are indicated and the associated probability is **p = 0.004. **(H-I)** Neutrophils from HD (n = 5), severe or critical COVID-19 patients (n = 5) were labeled with a mouse monoclonal anti-S100A8 antibody and with DAPI to visualize nuclei using the Widefield Zeiss Observer Z1 microscope (magnification x 100). **(H)** Representative experiment of S100A8 immunofluorescent staining showing its cytosolic localization (red) and the nuclei (blue). **(I)** Quantification of the S100A8 immunofluorescent staining. Photos were captured from at least 3 different fields for a given donor and MFI was analyzed in each field. Graph symbols represent individual donors. Data were analyzed by one way-ANOVA followed by Tukey or Dunnett post-test or by Student’s t-test for two groups and shown as mean ± SEM (**p<0.01; ***p<0.001).

### Neutrophils from COVID-19 patients present altered phenotype related with activation and immaturity

Neutrophils from both severe and critical COVID-19 patients showed increased levels of adhesion molecules such as integrin CD11b, CD66b (CEACAM8), the receptor-linked protein tyrosine phosphatase CD45, the metabolic marker lectin-like oxidized low-density lipoprotein receptor-1 (LOX-1) and programmed cell death protein ligand 1 (PDL-1), as well as decreased levels of CD62L (L-selectin) strongly suggesting that neutrophil from severe and critical COVID-19 patients were activated (Figure 5A). Moreover, a decreased expression of neutral endopeptidase CD10 was observed in COVID-19 patients compared to HD. Frequency of neutrophils expressing CD10 was decreased while frequency of those expressing LOX-1, PDL-1, Siglec-3 CD33 and CD114 was increased compared to HD (Supplemental Figure 4). Non-supervised analysis of neutrophils from HD, severe and critical COVID-19 patients showed an overall decrease in CD62L and increase in CD11b expression in the critical COVID-19 group (Figure 5B). We identified an enrichment of a neutrophil subset in critical COVID-19 patients, composed of very mature neutrophils (CD10^high^), with a particularly high activation state (CD11b^high^, CD66b^high^ and a LOX-1^high^ and CD114^high^ expression. The expansion of this hyperactivated subset of neutrophils is consistent with the overwhelming inflammatory state of critical COVID-19 patients.

**Figure 5.**
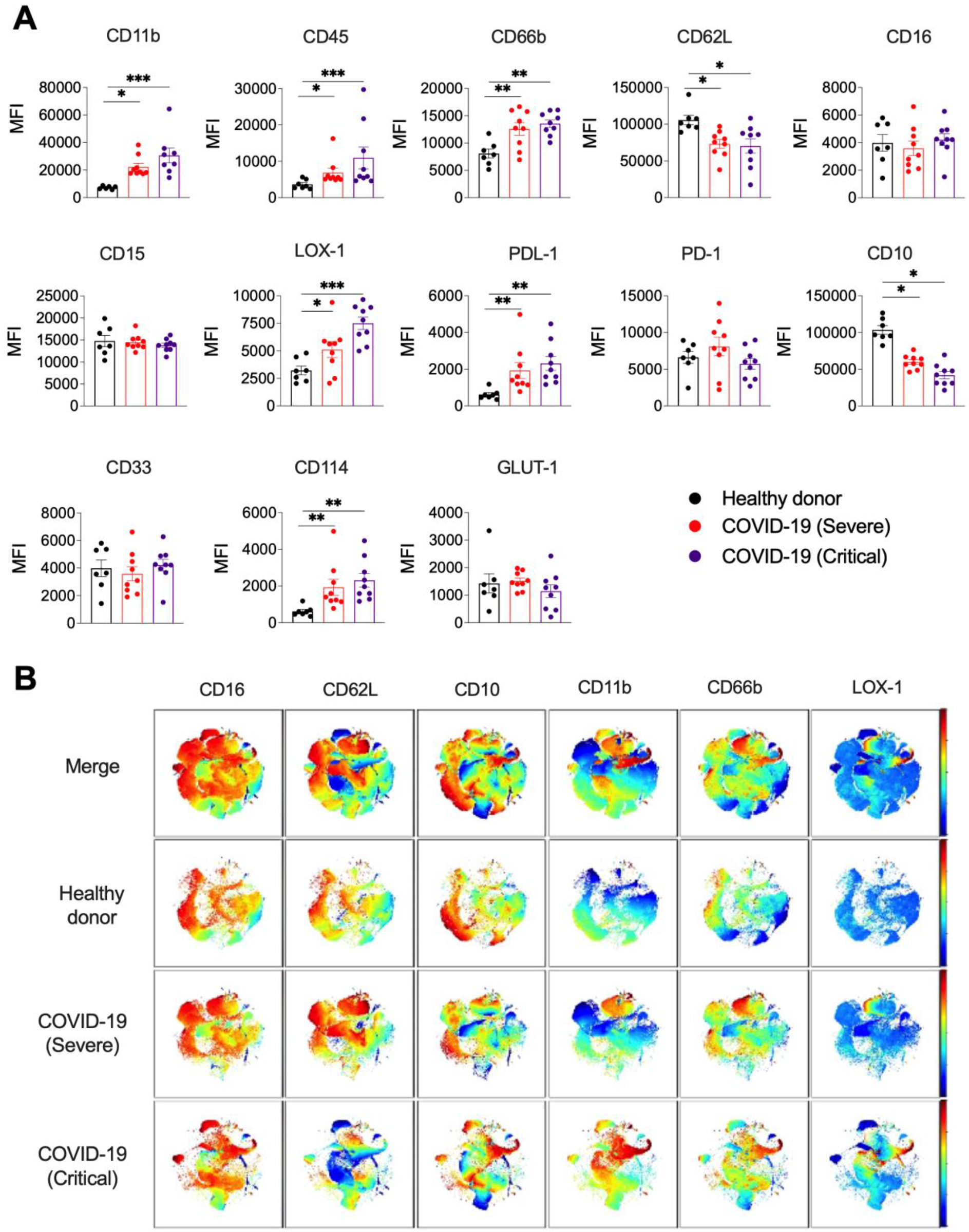
Neutrophils in COVID-19 patients present an altered phenotype linked to activation and immaturity. **(A)** Whole blood from HD (n = 7), severe (n = 10) and critical (n = 10) COVID-19 patients were used for leukocytes staining and the assessment of 13 surface markers was performed by spectral flow cytometry on a Cytek Aurora cytometer. Analysis was conducted in CD15^+^ cells using the gate strategies shown in Supplemental Figure 1. Results are expressed by MFI for each marker. **(B)** t-Distributed Stochastic Neighbor Embedding (t-SNE) plots showing neutrophil heterogeneity. Analysis was performed in CD15^+^ cells (n = 30000 down sampled) comparing HD (n = 5), severe (n = 5) and critical (n = 5) COVID-19 patients. Data were analyzed by one way-ANOVA followed by Tukey or Dunnett post-test or by Student’s t-test for two groups and shown as mean ± SEM (**p<0.01; ***p<0.001).

### PCNA-S100A8 cytosolic interaction is increased in an hyperactivated subset of CD16^high^/CD62L^low^ neutrophils in COVID-19

Owing to this neutrophil heterogeneity in COVID-19 patients, we next reasoned that the PCNA-S100A8 interaction may be associated with a particular subset of neutrophils endowed with specific functions. To address this issue, we evaluated the phenotype of neutrophils using a previously described gate strategy^43^ according to the CD16/CD62L staining pattern. In HD, most neutrophils presented high levels of CD16 and CD62L (CD16^high^/CD62L^high^), representing the conventional phenotype while the other subsets (CD16^high^/CD62L^low^, CD16^low^/CD62L^low^ and CD16^low^/CD62L^high^) were approximately 1%. In severe and critical COVID-19, we observed an expansion of neutrophil subsets presenting CD16^high^/CD62L^low^ (2), CD16^low^/CD62L^low^ (3) and CD16^low^/CD62L^high^ (4) characteristics and a decrease in the conventional CD16^high^/CD62L^high^ neutrophils was found (Figure 6A-B).

**Figure 6.**
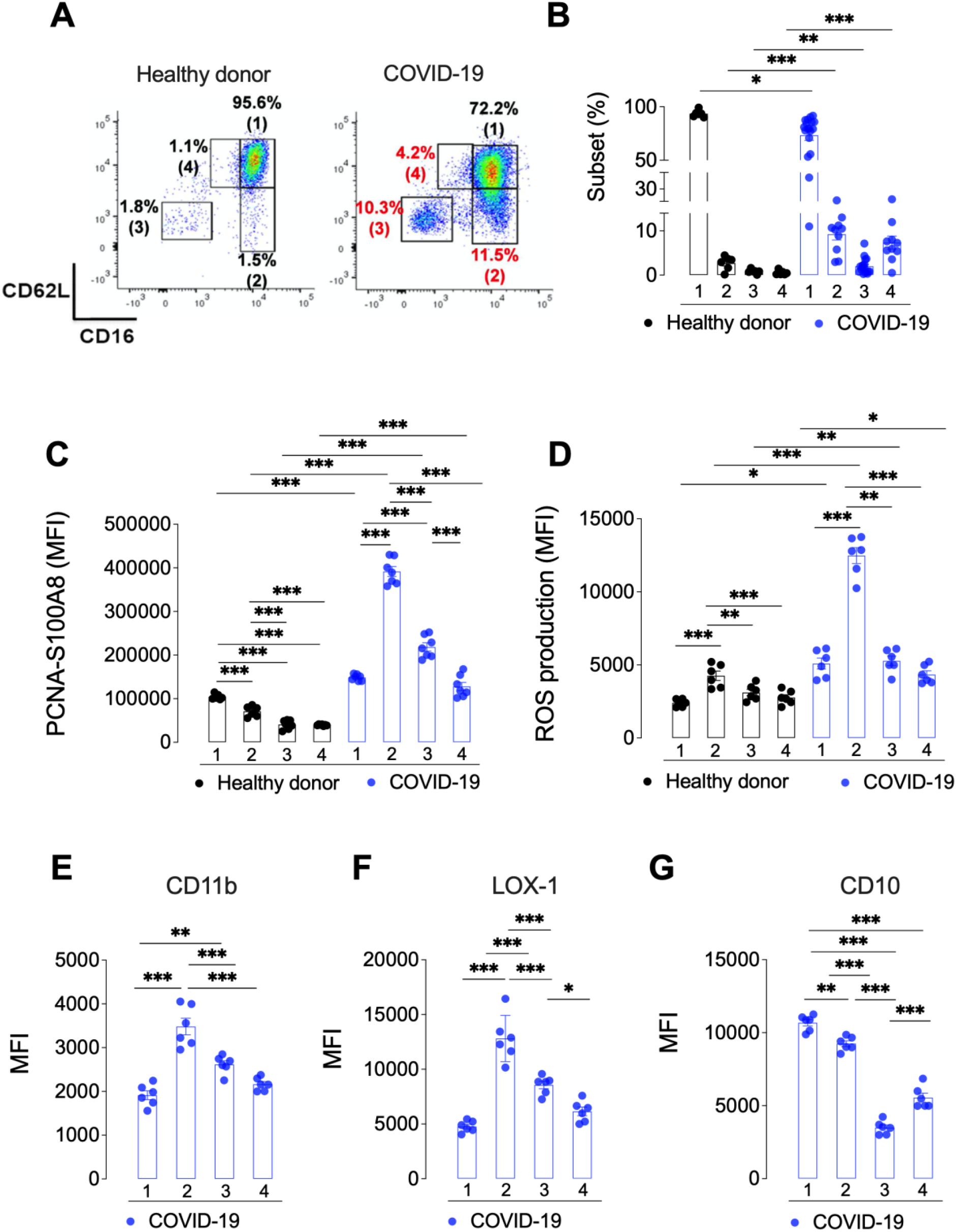
PCNA-S100A8 interaction is increased in an hyperactivated subset of neutrophils in COVID-19. **(A)** Representative flow cytometry dot plot showing the distinct neutrophil subsets according to CD16 and CD62L membrane labelling in neutrophils (CD15^+^ cells). Different subsets are observed in the HD compared to the COVID-19 patient. The percentages of positive cells (%) in the different CD16/CD62L neutrophil subsets that display CD16^high^/CD62L^high^ (subset 1), CD16^high^/CD62L^low^ (subset 2), CD16^low^/CD62L^low^ (subset 3) and CD16^low^/CD62L^high^ (subset 4) surface staining are presented. **(B)** Percentage of neutrophils in each CD16/CD62L subset. Neutrophil phenotyping was performed in blood from HD (n = 7), severe (n = 10) and critical (n=18) COVID-19 patients according to the gate strategy described in Supplemental Figure 1. **(C)** Flow cytometry quantification of the PCNA-S100A8 interaction by proximity-induced fluorescence using Duolink^®^ in the different neutrophil subsets (1-4) according to CD16/CD62L surface expression. **(D)** Measurement of ROS production using dichlorofluorescin diacetate (DCFDA) labelling in the different neutrophil subsets (1-4) according to CD16/CD62L surface expression. DCFDA-loaded neutrophils were next fixed and labeled for CD16 and CD62L surface expression. The expression of the surface markers **(E)** CD11b, (F) LOX-1 and **(G)** CD10 were also assessed in CD16/CD62L subset of neutrophils enriched in severe and critical COVID-19. Samples were acquired in a Cytek Aurora. Graph symbols represent individual donors. Data were analyzed by one way-ANOVA followed by Tukey or Dunnett post-test or by Student’s t-test for two groups and shown as mean ± SEM (**p<0.01; ***p<0.001).

Next, a Duolink proximity ligation assay was performed in combination with CD16/CD62L surface marker immunolabelling and was analyzed by flow cytometry. In HD, we noticed that the PCNA-S100A8 interaction was more intense in the conventional CD16^high^/CD62L^high^ subset (1) compared to the other subsets. By contrast, in severe and critical COVID-19, the PCNA-S100A8 interaction was more intense in the highly activated cells (CD16^high^/CD62L^low^) (2) than in the conventional CD16^high^/CD62L^high^ subset, strongly suggesting that the mechanisms leading to PCNA-S100A8 interaction could be different between HD and COVID-19 patients (Figure 6C). This was further corroborated by the measurement of ROS production after DCFCH staining combined with surface marker labelling analyzed by flow cytometry. Remarkably, at the basal state, every neutrophil subset from COVID-19 patients produced more ROS than their counterparts in HD (Figure 6D). This was also observed after PMA or opsonized zymosan stimulation (Supplemental Figure 5) thereby confirming the sustained NADPH-oxidase-dependent ROS production. Nonetheless, the ability of each subset to produce ROS in relation with the PCNA-S100A8 interaction seems to be different between HD and COVID-19 patients. In HD neutrophils, despite a lower level of PCNA-S100A8 interaction the CD16^high^/CD62L^low^ (2) subset (which represents less than 2% of the neutrophils) ROS production is higher in this subset compared to the conventional CD16^high^/CD62L^high^ (1) subset. In contrast, in COVID-19 neutrophils, the highest ROS production is observed in the CD16^high^/CD62L^low^ (2) subset which also has the highest level of PCNA-S100A8 interaction strongly suggesting that the PCNA-S100A8 interaction differentially affects ROS production in HD versus COVID-19 neutrophils. Furthermore, in COVID-19, the CD16^high^/CD62L^low^ subset (2) had the largest expression of CD11b (Figure 6E) and LOX-1 (Figure 6F), while showing decreased levels of CD10 when compared to CD16^high^/CD62L^high^ neutrophils (Figure 6G). In conclusion, the PCNA-S100A8 interaction is related to the production of large amounts of ROS and took place in a specific subset of mature and hyperactivated neutrophils from patients with severe COVID-19.

## Discussion

An excessive inflammatory response and dysregulated neutrophil phenotype are hallmarks of COVID-19 infection^15,21,44–51^ but the molecular defects in the control of neutrophil functions are still unknown. PCNA is a highly conserved scaffolding protein originally described in the nuclei of proliferating cells as a maestro of the replication.^24,25^ Indeed, the latter process requires a myriad of proteins that should interact together in a tightly regulated manner to maintain genome integrity.^52,53^ Likewise, neutrophil cytosolic PCNA orchestrates a protein scaffold that controls neutrophil functions including apoptosis, NADPH oxidase activity, and metabolism.^30,31^ In addition, PCNA is a key regulator of neutrophil survival through sequestration of procaspases to prevent their activation. During apoptosis, PCNA is degraded by the proteasome, thereby allowing the release of active caspases.^22^ In addition, the PCNA scaffold is extremely versatile in response to neutrophil activation but until now all the functions of cytosolic PCNA are poorly understood.

We first demonstrated that PCNA protein levels were increased in the cytosol of neutrophils according to COVID-19 severity. Of note, publicly available databases^16,41^ have not reported any increased PCNA mRNA in neutrophils from COVID-19 patients, whereas a proteomic analysis performed on neutrophil lysates has reported that PCNA was significantly upregulated in severe COVID-19 patients.^15^ This lack of transcriptional regulation of PCNA in neutrophils is in keeping with our previous finding showing that PCNA expression is significantly increased in neutrophils from donors treated with G-CSF or from patients with chronic inflammatory disease without any mRNA modulation.^22^ Notably, in COVID-19 patients, cytosolic PCNA levels were correlated with circulating markers of neutrophil-derived mediators such as MPO-associated DNA (NETs), neutrophil elastase, or more importantly, with seric calprotectin (S100A8/A9) concentration, this latter being a pronostic marker of the severity of the COVID-19.^16^ Indeed, previous studies report its altered mRNA transcription in both circulating and respiratory tract infiltrated neutrophils.^16,41^ We show for the first time that cytosolic S100A8 levels were significantly increased in cytosols from COVID-19 neutrophils and that they correlated with those of PCNA. Moreover, PCNA cytosolic levels were highly sustained even under physiological apoptosis conditions in neutrophils from severe/critical COVID-19 patients, thus maintaining their survival. S100A8, mainly localized in neutrophil cytosols and released in culture supernatants in association with NETs^54^, acts as a pro-survival factor suppressing neutrophil apoptosis via a TLR4 and NF-κB dependent mechanism.^55,56^ Hence, we concluded that both PCNA and S100A8 act together, favoring neutrophil survival and promoting their persistence in inflammation sites during serious cases of COVID-19 thereby contributing to the worsening of critical COVID-19 patients.

We previously demonstrated that PCNA is directly associated with the phox homology domain of the p47^phox^, a key subunit of NADPH-oxidase to regulate ROS production in human neutrophils.^31^ At basal state, this association prevents NADPH oxidase activation, whereas upon activation, PCNA acts as a chaperone by favoring its assembly and then dissociates from the p47^phox^ protein. Here, we showed that the association between PCNA and p47^phox^ was disrupted in neutrophils from COVID-19 patients suggesting a defect in this gate keeper function of PCNA on NADPH oxidase and presumably, a modified PCNA scaffold. S100A8/S100A9 is also implicated in the positive modulation of the NADPH oxidase via molecular interactions with p67^phox^, Rac-2,^32^ and cytochrome b_558_,^33^ respectively the cytosolic and membrane associated components of the enzyme. We provide evidence that during COVID-19 PCNA was preferentially associated with S100A8, a process dramatically inhibited by the PCNA scaffold inhibitor T2AA, thereby suggesting that the PCNA-S100A8 cytosolic partnership may potentiate NADPH-oxidase activity, reinforcing the concept of PCNA as a key oxidative burst modulator in COVID-19. Unraveling the molecular details of this PCNA-S100A8 interaction will require further structural biology experiments to determine the precise PCNA interacting domain in S100A8.^57,58^ Nonetheless, our findings may provide the basis of a PCNA targeted therapy to specifically dampen this overactivation of NADPH oxidase. Using phenotyping analysis combined with single cell analysis a robust enrichment of CD16^low^ neutrophil subsets characterized by immaturity^59^ and activation (CD62L^low^) has been observed in severe COVID-19 patients.^60–62^ Those cells are also characterized by higher levels of CD66b, LOX-1, and CD24, which are associated with early stages of neutrophil development in COVID-19.^61,63^ In another study, LOX-1-expressing immature neutrophils are suggested to identify critically ill COVID-19 patients at risk of thrombotic complications.^64^ Regarding neutrophil phenotyping, we obtained similar results indicating that COVID-19 neutrophils are basally activated with altered levels of CD11b, CD45, CD114 and CD62L, and characteristics of immaturity and immunosuppression indicated by LOX-1, CD10 and CD16 assessment. Additionally, we found neutrophils from COVID-19 express higher surface levels of PDL-1 and CD33 (Siglec-3), markers associated with immunosuppression.^21,61,65–67^ Determination of the CD16/CD62L subsets in various conditions has proven to be a reliable method to approach the phenotypic and functional analysis of neutrophil diversity^43,59^. Neutrophils at homeostasis present very uniform expression of CD16 and CD62L (CD16^high^/CD62L^high^) that progressively changes by the appearance of two other subpopulations after intravenous LPS challenge, CD16^low^/CD62L^high^ and CD16^high^/CD62L^low^.^43^ According to a morphological and functional analysis of those two subsets, CD16^low^/CD62L^high^ have banded nuclei indicating immaturity and show increased resistance to physiological apoptosis.^43^ This subset is characteristic of acute inflammation observed in LPS exposure, bacterial sepsis, acute trauma^68^. In contrast, CD16^high^/CD62L^low^ neutrophils present hyper segmented nuclei, high expression of activation markers, such as CD11b, CD11c and CD54 (ICAM), increased NADPH-oxidase activity, and T cell suppression effects.^43^ Although this CD16^high^/CD62L^low^ subset can be detected in some acute condition such as LPS injection, it seems to be a hallmark of a chronic inflammatory challenge such as trauma or COVID-19. Notably, in the COVID-19 patients returning from ICU patients, the sorted CD16^high^/CD62L^low^ subset had a hypersegmented nuclear morphology and show an increased CD11b expression suggesting an activated state. Accordingly, we also observed that in severe and critical COVID-19 neutrophils were CD16^high^/CD62L^high^, CD16^high^/CD62L^low^ and CD16^low^/CD62L^high^, but also as CD16^low^/CD62L^low^. This last population may represent an immature neutrophil subset, marked by low levels of CD10. In addition, COVID-19 critical patients present higher plasmatic levels of G-CSF, accompanied by an emergency myelopoiesis and early neutrophil release from the bone marrow,^67–70^ explaining the expansion of CD16^low^ immature neutrophils.^69^ In agreement, we observed that severe and critical patients showed increased percentages of CD114^+^ neutrophils, the G-CSF receptor.

Most importantly, our study provides new insights about the impact of the PCNA scaffold on neutrophil heterogeneity and functions, therefore proposing that PCNA may orchestrate dedicated functions in specific subsets. Indeed, PCNA-S100A8 interaction was detected in conventional neutrophils, which display a very homogeneous CD16^high^/CD62L^high^ phenotype. By contrast, in neutrophils from severe and critical COVID-19 patients, we observed a dramatic increase in the intensity of this interaction but also a radical switch to the CD16^high^/CD62L^low^ subset in which this interaction was the most intense. Pertinently, this PCNA-S100A8^high^ subset had increased ROS production, and increased CD11b and LOX1 expression as compared to the conventional CD16^high^/CD62L^high^ subset. Nonetheless, this PCNA-S100A8 ^high^ subset is not immature since no decreased CD10 expression was observed compared to the main CD16^high^/CD62L^high^ subset. Further investigations are required to decipher the immunomodulatory functions of this PCNA-S100A8^high^ subset^70^ which accounts for more than 10% of the circulating neutrophils in COVID-19 patients and which may be considered as a pathogenic and pro-inflammatory subpopulation. Accordingly, inhibiting the molecular interaction between PCNA and S100A8 would provide a unique opportunity to target this pro-inflammatory subset of neutrophils without affecting the general neutrophil population, thereby avoiding neutropenia to maintain an efficient anti-infectious defense.

In conclusion, we propose that the increased neutrophil cytosolic PCNA is involved in the profound immune dysregulation observed in patients with severe COVID-19. Decoding the entire scaffold would certainly provide new insights into the molecular and functional dysregulation of neutrophils in patients with COVID-19.

## Materials and methods

### Human blood samples

Individuals hospitalized due to COVID-19 from April 2020 to January 2022 were recruited in Paris, at the Cochin Hospital from the Internal Medicine department (authorization #2019-3677), the Respiratory medicine department (authorization #2020 #A02700-39) and from the Intensive Care Medicine department (authorization #2018 #A01934-51) and at Bichat Hospital from the Intensive Care Medicine department (authorization #2020-715). This study was approved by the Hospital Ethics Committee, according to the Declaration of Helsinki. Each patient gave written informed consent and was classified into different clinical severity groups (Table 1) depending on a multifactorial score taking into account oxygen requirement.^34–36^ Blood from COVID-19 patients and from healthy donors (HD) obtained from the *Etablissement Francais du Sang* was collected in EDTA-vacuum or in dry tubes for serum collection. Neutrophils were isolated in LPS-free dextran sedimentation and Ficoll (Histopaque-1077^®^, Sigma-Aldrich) centrifugation, as previously described.^22^

**Table 1.** Clinical and biological characteristics of patients

|  | COVID-19 |  |  |
| --- | --- | --- | --- |
|  | Moderate<br>(n=15) | Severe<br>(n = 21) | Critical<br>(n19) |
| Male sex (n/%) | 8 / 53.3% | 14 / 66.7% | 16 / 84.2% |
| Female sex (n/%) | 7 / 46.7% | 7 / 33.3% | 3 / 15.8% |
| Age* (years) | 76.9 ± 3.3 (42-89) | 68.7 ± 11.3 (44-92) | 64.6 ± 2,4 (48-77) |
| Leukocytes** (/μl) | 5910.7± 650 | 7367.5±2180.5 | 9326.5±1110 |
| Neutrophils** (/μL) | 4206.43±665.2 | 5971.4±2150.3 | 8135.26±1123.5 |
| CRP** (mg/dL) | NA | 46.3±13 | 90.3±18.7 |
| D-Dimer** (ng/ml) | 1212±253 | 1444±910 | 3293±1295 |
| Oxygenotherapy at inclusion (n) | 6 | 21 | 17 |
| Oxygen flow < 3L/mn (n) | 5 | 2 | 2 |
| Oxygen flow > 3L/mn (n) | 1 | 10 | 10 |
| Nasal high flow oxygen (n, (%)) | 0 | 9 (42.9%) | 5 (29.4%) |
| Need for nasal high flow oxygen during hospitalization (n, (%)) | 4 | 15 (71.4%) | 13 (68.4%) |
| Systemic corticotherapy (n) | 6 | 20 | 17 |
| Complete hospitalization duration (Days, Median (IQR)) | 16±2.5 |  | 15.7±3 |
| Death (n) | 0 | 1 | 7 |

\*Results are expressed as mean ± SEM (min-max); \*\*Results are expressed as mean ± SEM. NA: not available.
Moderate patients either do not require oxygen or require a flow of less than 3 L/min. Severe patients require COVID-19 oxygen flow between or over 3 L/min. Critically ill patients require more than 60% high flow oxygen and ventilation and have been recruited in the Intensive Care Unit (ICU).

### Quantification of circulating neutrophil-derived activation markers

Circulating neutrophil elastase (ThermoFisher Scientific), calprotectin (S100A8/A9) (Biotechne) and Neutrophil Extracellular Traps (NETs) were measured by ELISA as previously described^37^.

### Neutrophil phenotype assessment using spectral flow cytometry

Blood neutrophils from HD or COVID-19 patients were labelled with the following antibodies: CD66b, CD15, CD62L, CD16, CD11b, CXCR2, PD-1, CD45, CD10, CD33, CD114, PDL-1, GLUT-1, LOX-1 or their respective isotypes (Table 3). Samples were acquired in a Cytek Aurora cytometer (Cytek Biosciences, Fremont, CA, USA). Data were analyzed using FlowJo v10.7.1 software (FlowJo LLC) (Supplemental Figure 1). Neutrophil phenotype was also evaluated by unsupervised analysis using the ti-SNE dimensional reduction algorithm. A sample of 30000 neutrophils (CD15^+^ cells) from each individual were normalized and pulled down. Then, cells were grouped and subjected to the algorithm to form homogeneous clusters based on CD16, CD62L, CD10, CD11b, CD66b and LOX-1 surface expression using the CytoBank platform (Beckman Coulter Life Sciences, California).

**Table 3.** Antibodies used for the phenotyping of neutrophils by flow cytometry.

| Surface marker | clone | Commercial source |
| --- | --- | --- |
| CD66b | G10F5 | BioLegend |
| CD15 | W6D3 | BD Biosciences |
| CD62L | DREG-56 | BioLegend |
| CD16 | 3G8 | BioLegend |
| CD11b | ICRF44 | BioLegend |
| CXCR2 | 5E8 | BioLegend |
| PD-1 | EH12.1 | BD Biosciences |
| CD45 | 2D1 | BioLegend |
| CD10 | HI10a | BD Biosciences |
| CD33 | P67.6 | BD Biosciences |
| CD114 | LMM741 | BD Biosciences |
| PDL-1 | MH3 | BioLegend |
| GLUT-1 | 1418G | R&D Systems |
| LOX-1 | 15C4 | BioLegend |

### Assessment of physiological apoptosis

Neutrophils (2 × 10^6^ cells/mL in RPMI-1640-10% FBS) were incubated overnight (16h, 37°C, 5% CO_2_) and then labeled with annexin-V (Miltenyi Biotec) and 7-AAD (BD Biosciences) as previously described.^22^ To assess mitochondrial integrity, neutrophils were stained with 1 μM of 3,3’-dihexyloxacarbocyanine iodide (DiOC6), a fluorescent dye selective for live cell mitochondria^38,39^ (ThermoFisher). Data acquisition was performed using BD-Accuri™-C6Plus, analyzed by Cflow Plus software.

### Evaluation of NADPH-oxidase-dependent ROS production

Isolated neutrophils (0.1 × 10^6^ cells) were suspended in 0.1 mL HBSS with or without T2AA (Sigma-Aldrich) at indicated concentrations for 1 h (37°C, 5% CO_2_). Neutrophils were then stimulated with or without PMA (Sigma-Aldrich; 0.1 μg/ml), IgG- and complement proteins-opsonized zymosan (0.5 mg/mL), or N-Formyl-methionine-leucyl-phenylalanine (f-MLF) (Sigma-Aldrich; 10 μM) as previously described (Ohayon et al 2019). Chemiluminescence was recorded after addition of luminol (Sigma-Aldrich; 10 μM) at 37°C in a TRISTAR luminometer (Bertold, Wild bad Germany) over time. The data were expressed as integrated total counts, processed by the microWim software and analyzed using Prism 9.3 software. In order to characterize the membrane phenotype of ROS-producing neutrophils, cells were first loaded with 10 μM of cell-permeant 2’,7’-dichlorodihydrofluorescein diacetate (DCF-DA, Thermo Fisher) and were stimulated with or without PMA or opsonized zymosan and were next stained with a mix of anti-human CD16 and CD62L to determine neutrophil subpopulations. Samples were acquired in a Cytek Aurora (Cytek Biosciences, Fremont, CA, USA) cytometer and approximately 100 000 gated events were used in each analysis (Supplemental Figure 1). Data were analyzed using FlowJo v10.7.1 software (FlowJo LLC).

### Western blot analysis of PCNA and S100A8

Neutrophil cytosol was obtained by sonication in a hypotonic HEPES buffer (50 mM) supplemented with inhibitors (4 mM PMSF, 400 μM leupeptin, 400 μM pepstatin, 1 mM orthovanadate, 1 mM EGTA, and 1 mM EDTA) and DTT (1 mM), with a Soniprep 150 Plus sonicator (1 Hz for 10 s) as previously described.^40^ Alternatively, neutrophils were lysed in the sonication buffer supplemented with 1% NP40 detergent to obtain a whole cell lysate. Protein concentration was measured using the BCA kit (Pierce). Samples diluted into Laemmli buffer (30 μg per well) were subjected to SDS-PAGE, transferred to polyvinylidene difluoride (PVDF) membrane (PerkinElmer Life Sciences). After blocking with Tris-buffered saline (TBS) containing 5% nonfat dry milk, membranes were probed with mouse monoclonal anti-PCNA (PC10; 1:1000 dilution), goat polyclonal anti-S100A8 (Everest, EB11513; 1:1000 dilution), goat polyclonal anti-actin (Sigma A2066; 1:1000 dilution) antibodies overnight at 4°C. Secondary horseradish peroxidase-conjugated-linked anti-rabbit IgG (Jackson Lab, 111-035-003; 1:5000), anti-mouse IgG (Jackson Lab, 115-036-006; 1:2000) or anti-goat IgG (Santa Cruz biotechnology, sc-2033; 1:4000) antibodies were applied 1h at room temperature as previously described (Witko-Sarsat et al 2010). Immunoreactivity was visualized using enhanced chemiluminescence reagents (Amersham Biosciences). Images were acquired and quantified using a Fusion FX7 system imaging (Vilbert Lourmat).

### PCNA and S100A8 detection using indirect immunofluorescence or the Duolink^®^ proximity ligation assay (PLA)

Cells were fixed in 2% formaldehyde/PBS for 20 min on ice and permeabilized with 0.25% Triton X-100 for 5 min at room temperature, followed by ice-cold methanol for 10 min, incubated with the rabbit polyclonal anti-PCNA (Ab5; 1:25) or the mouse monoclonal anti-S100A8 (2H2; 1:100) antibodies for 45 min. PCNA incubation was followed by biotinylated anti-rabbit IgG (Dako; 1:200) for 30 min, and then by streptavidin-coupled Alexa Fluor 555 (Invitrogen; 1:1000) for 30 min. S100A8 incubation was followed by anti-mouse IgG Alexa Fluor 488 (Invitrogen; 1:100) for 30 min. Nuclei were stained with DAPI containing mounting media (Sigma Aldrich) as previously described.^22^ Red fluorescence was observed with a 488–560 nm-long-pass emission filter under 543 nm laser illumination. Images were processed using ImageJ software version 1.53J (National Institutes of Health) and fluorescence normalized to DAPI fluorescence as previously described

For Duolink^®^ PLA assay, neutrophils (0.3 × 10^6^) were processed as described by the manufacturer using a mix of anti-PCNA (Ab5; 1:100) and anti-S100A8 (2H2; 1:100) as primary antibodies either on cytospins or in suspension for microscopic or flow cytometry analysis, respectively. Detection was achieved if distance between the two targeted epitopes are up to 40 nm. Fluorescence was acquired under microscopy (Widefield Zeiss Observer Z1 and SR Leica SP8X STED Flim) or by FACS (Cytek Aurora) gating at least 100 000 neutrophils. Data were analyzed using ImageJ 1.53J and FlowJo v10.7.1 softwares, respectively.

### Analysis of PCNA-S100A8 interaction by surface plasmon resonance (SPR)

PCNA was obtained using *E. coli* vector expression and purification as previously described^30^. Interactions between PCNA and recombinant human (rh) S100 proteins (R&D systems, Biotechne, MN, USA) were studied by SPR on a Biacore T200 (GE Healthcare) designed to calculate affinity and kinetic parameters as previously described.^22,31^ Briefly PCNA was covalently immobilized on a dextran layer sensor chip (CM5) while the rh S100A8 (9876-S8), S100A9 (9254-S9) or S100A8/A9 (8226-S8) (1:1 complex) protein at the indicated concentrations (0.1–2 μM) were used as analytes in a running buffer as previously described. In all experiments, the specific binding signal was obtained by subtracting the background signal. For global fitting we used kinetic titration on the Biacore T200 system, referred to as multiple-cycle kinetics, to collect binding data for kinetic analysis. This method involves sequential injections of several analyte concentrations over the ligand immobilized on the sensor chip surface after a regeneration step between successive injections by 5 mM EDTA. The association (K_a_) and dissociation (K_d_) rate constants and K_D_ were calculated using BIAevaluation software. The simplest 1:1 Langmuir binding model was first tested. The heterogeneous ligand model was used as it significantly increased the statistical goodness-of-fit based on the *Chi*^*2*^ value as calculated by Biacore software. This model accounts for the presence of two sites that bind analyte independently of each other.

### Statistical analysis

Data are expressed as mean ± standard error of the mean (SEM) from (n) independent experiments performed on neutrophils isolated from different donors or patients and analyzed by one-way ANOVA followed by Dunnett’s or Tukey’s test. For paired comparisons, Student’s t-test was used. GraphPad Prism Software 9.0 was used for data processing (\**P* < 0.05; \*\**P* < 0.01; \*\*\**P* < 0.001 were considered statistically significant).

## Data Availability

All data produced in the present study are available upon reasonable request to the authors

## Online supplemental material

Supplemental Figure 1. Representative gate strategy used for neutrophils analysis by flow cytometry.

Supplemental Figure 2. Re-analysis of publicly available RNA-seq data of PCNA and S100A8. Expression of PCNA and S100A8/A9 genes in circulating and respiratory tract-infiltrated neutrophils from COVID-19 patients.

Supplemental Figure 3. Concentration-response curve of T2AA, a PCNA inhibitor, in the activation of NADPH-oxidase in neutrophils from healthy controls and critical COVID-19 patients.

Supplemental Figure 4. Phenotypic characterization of whole blood neutrophils in COVID-19 patients.

Supplemental Figure 5. Phenotypic characterization of low-density-neutrophils in the PBMCin COVID-19 patients.

## Author contributions

All named authors meet the general criteria for authorship of this manuscript and have given final approval for publication. ROF, LP, LT, SM, VK, KB, MA and VWS designed experiments and analyzed data; ROF, LP, VK, SM, ML, LT, TD, MS, ELR, EPL, PF, VG, LCM, FS, CR, MC performed experiments; MHN, AH, SCM, PRB, LM, CM, FP contributed with critical reagents/tools/clinical samples; ROF, LP, ML, LT and VWS wrote the manuscript.

## Acknowledgments

We are grateful for the excellent technical help provided by Giovanni Saracenti-Tasso to isolate neutrophils and to measure the respiratory burst, Thomas Guilbert (IMAGIC/Institut Cochin) and Nicolas Cagnard (Bioinformatics Core Platform/Imagine Institute) for the technical support. We thank Molly Ingersoll and Karen Aymonnier for review and feedback on the manuscript.

This study was supported by the Fondation pour la Recherche Médicale Grant EQU202003010155 (VWS) and the Investissements d’Avenir programme ANR-11-IDEX-0005-02, Sorbonne Paris Cite, LabEx INFLAMEX (VWS); the French National Research Agency (ANR) with the Grant RA-COVID-19 V5-COVINNATE (MHN); Grant DENDRISEPSIS ANR-17-CE15-0003 (FP); Grant APCOD ANR-17-CE15-0003-01 (FP); the Sao Paulo research Foundation Grant FAPESP-SCRIPPS 15/50387-4 (FS)

