## Supplementals for "Cytosolic PCNA interacts with S100A8 and controls an inflammatory subset of neutrophils in COVID-19"

**Supplemental Figures**

### Cytosolic proliferating cell nuclear antigen interacts with S100A8 and controls an inflammatory subset of neutrophils in COVID-19

Rodrigo de Oliveira Formiga, Lucie Pesenti, Maha Zohra Ladjemi, Philippe Frachet, Muriel Andrieu, Souganya Many, Vaarany Karunanithy, Karine Bailly, Théo Dhôte, Manon Castel, Christophe Rousseau, Marick Starick, Edroaldo Lummertz da Rocha, Emilia Puig Lombardi, Vanessa Granger, Sylvie Chollet-Martin, Luc De Chaisemartin, Luc Mouthon, Fernando Spiller, Anne Hosmalin, Margarita Hurtado-Nedelec, Clémence Martin, Frédéric Pène, Pierre-Regis Burgel, Léa Tourneur, Véronique Witko-Sarsat

**Supplemental Figure 1**

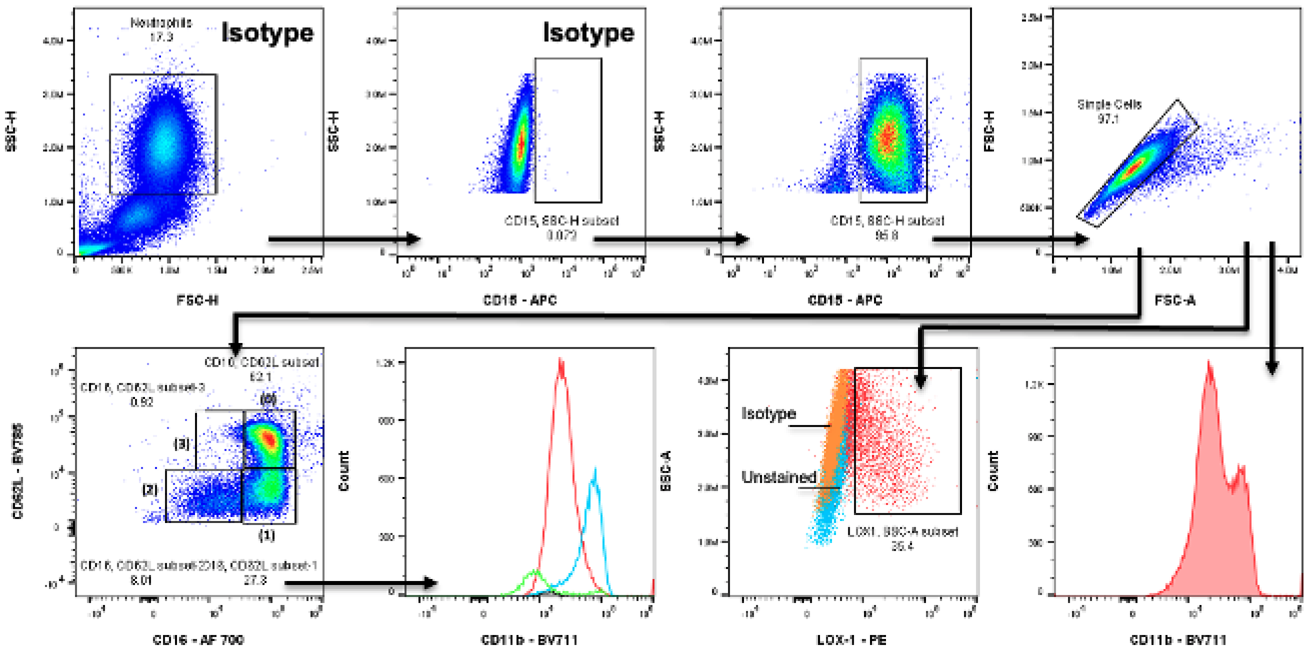


**Representative gate strategy used for neutrophils analysis by flow cytometry.**

For analysis of neutrophils stained in whole blood a classic side-scatter (SSC) versus forward-scatter (FSC) followed by the selection of $CD15^{+}$ cells according to the staining of the appropriate isotype control and exclusion of the double events (FSC-H vs. FSC-A) was carried out. Neutrophils were analyzed using the CD16 and CD62L staining pattern, the percentage numberof positive cells for a given marker or according to the median fluorescence intensity (MFI) value.

**Supplemental Figure 2**

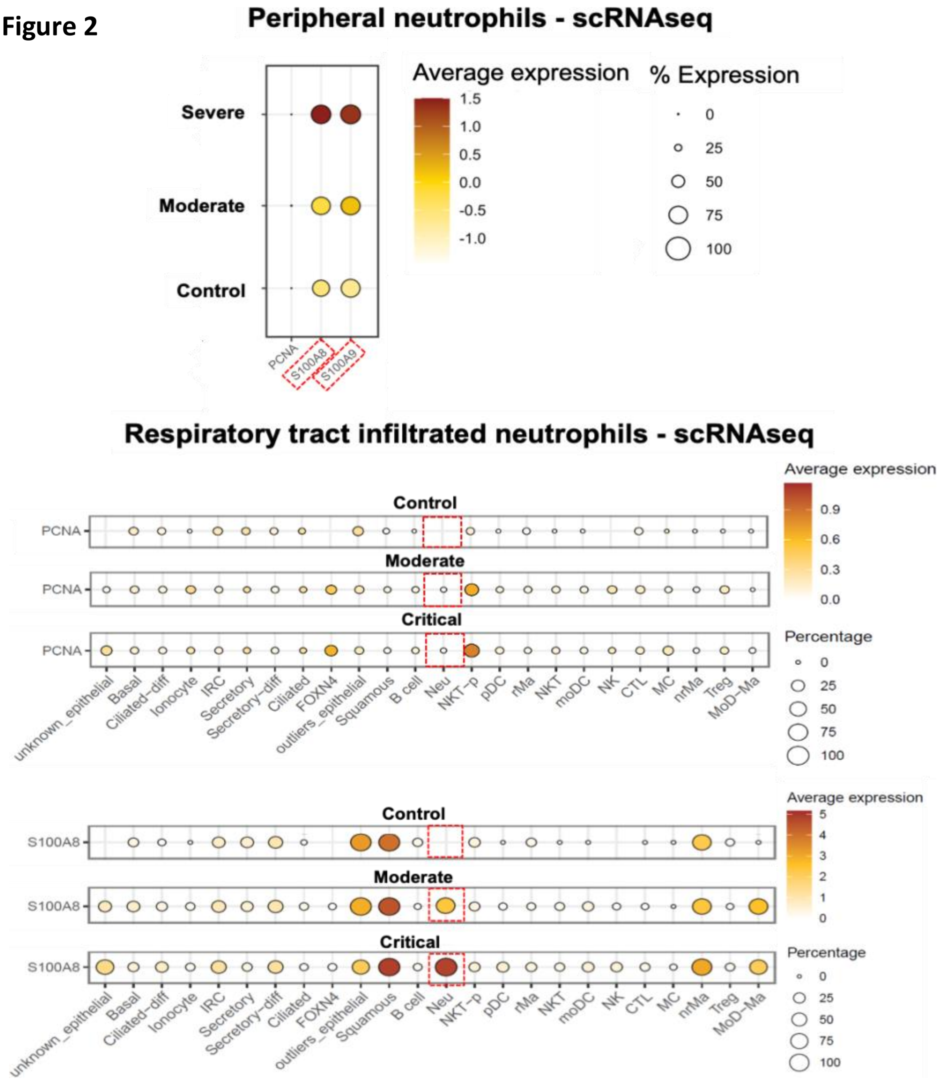


**Re-analysis of publicly available RNA-seq data of PCNA and S100A8. Expression of PCNA and S100A8/A9 genes in circulating and respiratory tract-infiltrated neutrophils from COVID-19 patients**.

The upper panel **(A)** shows the average and percentage of gene expression in whole blood isolated neutrophils from healthy donors(control), moderate or severe COVID-19 patients (*Silvin et al. Elevated Calprotectin and Abnormal Myeloid Cell Subsets Discriminate Severe from Mild COVID-19. Cell. 2020;182(6):1401-1418.e18*). The lower panels **(B-C)** present the average and percentage of gene expression in cell types isolated from nasopharyngeal/pharyngeal swabs samples from healthy donors (control), moderate or critical COVID-19 patients. The size of the circle is proportional to the percentage of cells expressing the reported genes at a normalized expressionlevel higher than one. Neu = neutrophils. (*Chua et al. COVID-19 severity correlates with airway epithelium-immune cell interactions identified by single-cell analysis. Nat Biotechnol. 2020 Aug;38(8):970-979*) **(D)** Identification of neutrophil population in nasopharynx swabs. UMAP (Uniform Manifold Approximation and Projection) analysis colored-coded by cell types in nasopharyngeal/pharyngeal swabs samples from severe COVID-19 patients. **(E-F)** Normalized expression overlaid on the UMAP spaces for **(E)** PCNA and **(F)** S100A8. This overlay provides evidence that PCNA mRNA expression is weak in neutrophils (identified in green) compared to its expression in other cell types including immune cells (NKT cells or in macrophages) or in epithelial cells. In contrast, S100A8 mRNA is strongly upregulated in neutrophils, macrophages, dendritic cells and epithelial cells. *(Chua et al. COVID-19 severity correlates with airway epithelium-immune cell interactions identified by single-cell analysis. Nat Biotechnol. 2020 Aug;38(8):970-97).*

**Supplemental Figure 3**

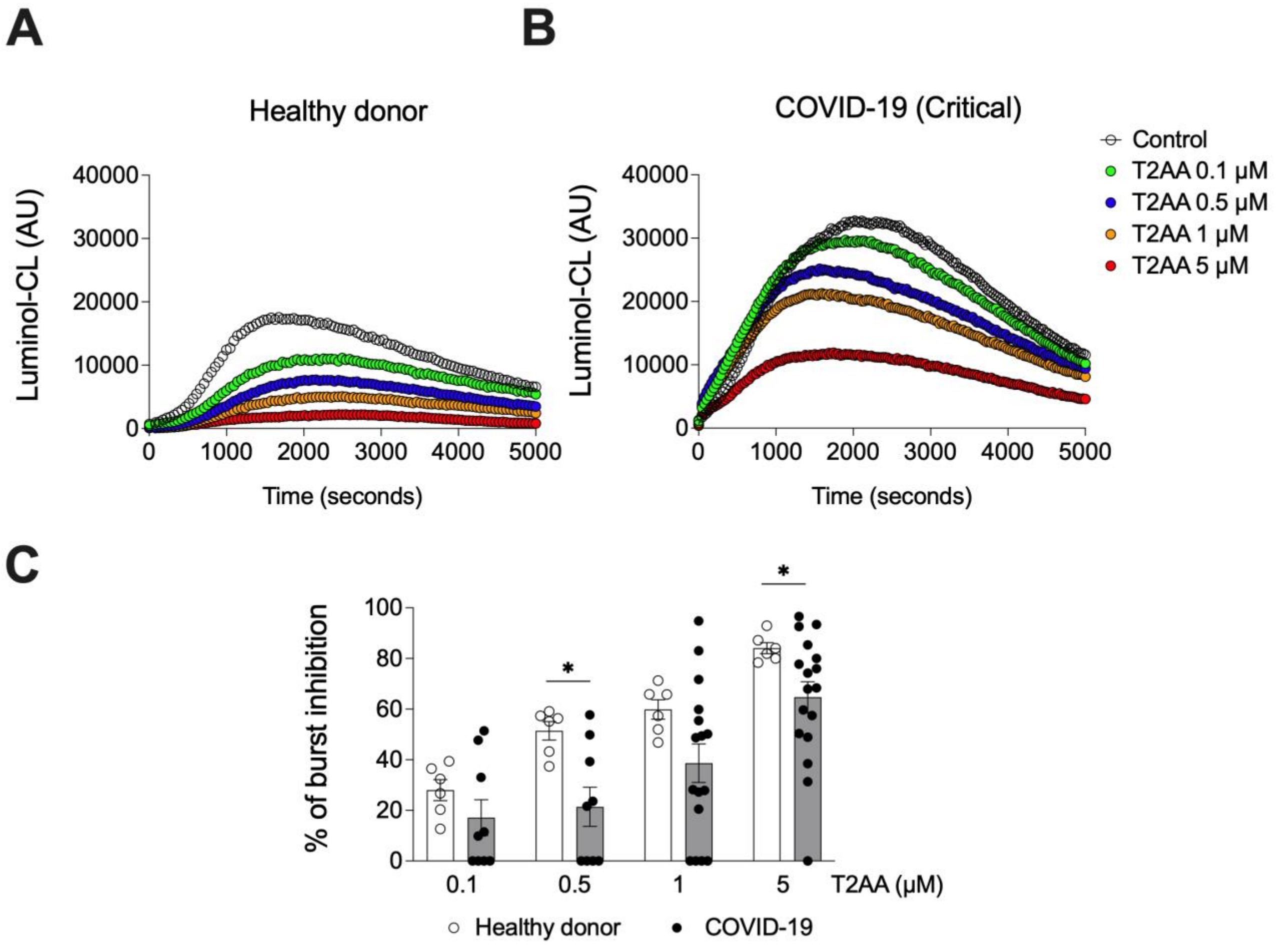


**Dose-response curve of the effect of T2AA, a PCNA inhibitor, on NADPH-oxidase-dependent ROS production in neutrophils.**

NADPH-oxidase activation was assessed by luminol-enhanced chemiluminescence (CL) that measured ROS production in isolated neutrophils from **(A)** healthy controls (n = 6) and **(B)** critical COVID-19 patients after stimulation by opsonized zymosan (0.5 mg/mL). These measurements were performed in the absence or in the presence of the PCNA inhibitor, T2AA at different concentration ranging from 0.1 to 5 µM. CL was recorded at 37°C in a Tristar luminometer over time and data were expressed as arbitrary units (AU). **(C)** Percentage of oxidative burst inhibition by T2AA under opsonized zymosan stimulation. T2AA percentage of inhibition under stimulation has been calculated for each concentration from healthy controls (n = 6) and critical COVID-19 patients (n=17). For instance, a 60% inhibition of opsonized zymosan-induced ROS production is observed at 1 and 5µM of T2AA in neutrophils from HD and COVID-19 patients, respectively. Data are presented as mean ± SEM. Data was analyzed by two way-ANOVA followed or by t test (*p<0.05; ***p<0.001).

**Supplemental Figure 4**

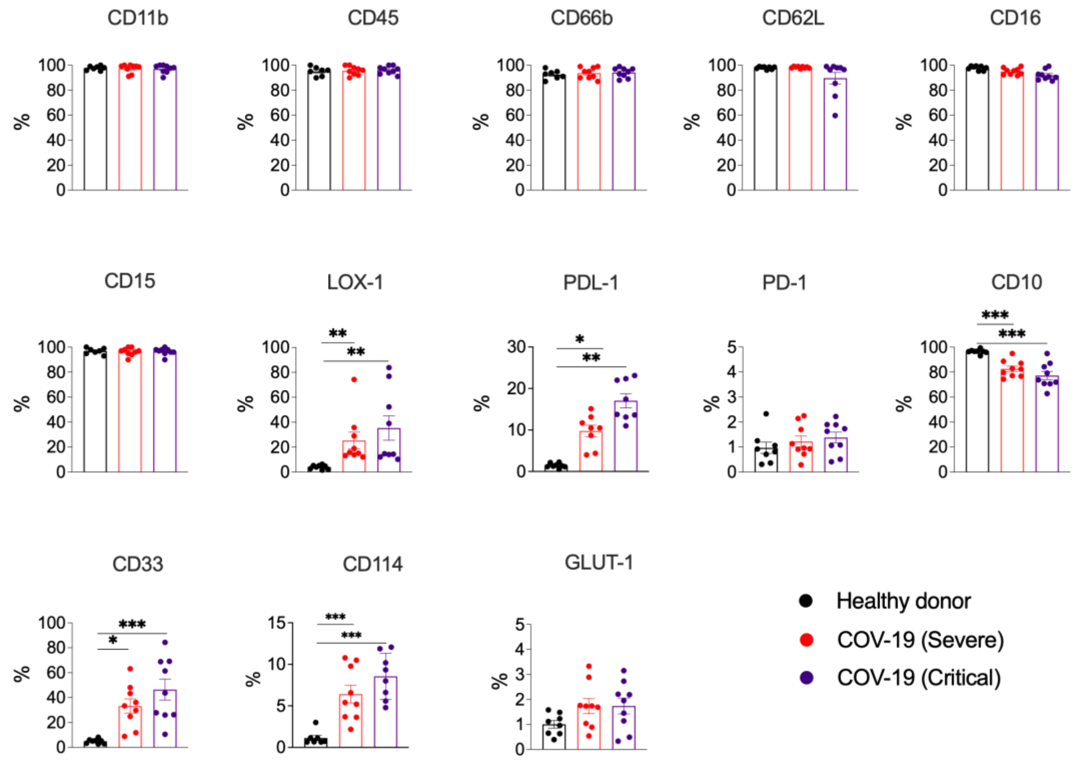


**Phenotypic characterization of whole blood neutrophils in COVID-19 patients.**

Total blood from healthy controls (n = 7), severe (n = 9) and critical (n = 9) COVID-19 patients were used for leukocytes staining and the assessment of the surface markers by spectral flow cytometry performed on a Cytek Aurora cytometer CD11b, CD45, CD66b, CD62L, CD16, CD15, LOX-1, PDL-1, PD-1, CD10, CD33, CD114 and GLUT-1. Results are expressed as percentage (%) in relation to the respective isotype control. Analysis was conducted in $CD15^{+}$ cells using the gate strategies shown in Supplemental Figure 1.

Data were analyzed by one way-ANOVA followed by Tukey or Dunnett post-test or by Student's t-test for two groups and shown as mean ± SEM (*p<0.05; **p<0.01; ***p<0.001). CD = cluster of differentiation; LOX-1 = lectin-like oxidized low-density lipoprotein receptor-1; PDL-1 = programmed cell death protein ligand 1; PD-1 = Programmed cell death protein 1; GLUT-1 = glucose transporter 1. SEM = standard error of the mean.

**Supplemental Figure 5**

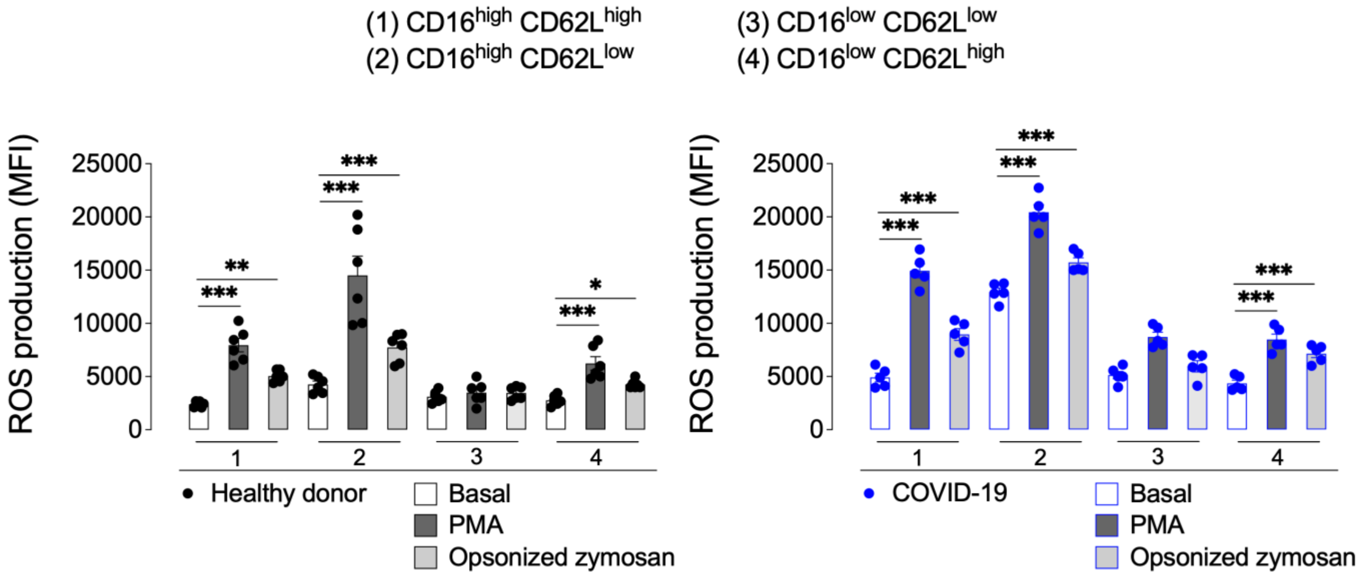


**Analysis of ROS production in each neutrophil subset according to their CD16/CD62L phenotype.**

In order to characterize the membrane phenotype of ROS-producing neutrophils, cells were first incubated with 10 µM of cell-permeant 2',7'-dichlorodihydrofluorescein diacetate (DCF-DA) and were next stimulated (or not) by PMA or opsonized zymosan for 10 min. Cells were fixed and stained with a mix of anti-human CD16 and CD62L antibodies to determine neutrophil subpopulations. Samples were acquired in a Cytek Aurora.
ROS production at basal state or after stimulation with PMA or opsonized zymosan in each neutrophil subset as defined above and is expressed as MFI in neutrophils from healthy donors (left panel) and from COVID patients (right panel).

Graph symbols represent individual donors. Data were analyzed by one way-ANOVA followed by Tukey or Dunnett post-test or by Student's t-test for two groups and shown as mean ± SEM (**p<0.01; ***p<0.001). MFI = median fluorescence intensity
